# THE LOW-HARM SCORE FOR PREDICTING MORTALITY IN PATIENTS DIAGNOSED WITH COVID-19: A MULTICENTRIC VALIDATION STUDY

**DOI:** 10.1101/2020.05.26.20111120

**Authors:** Adrian Soto-Mota, Braulio A. Marfil-Garza, Erick Martínez Rodríguez, José Omar Barreto Rodríguez, Alicia Estela López Romo, Paolo Alberti Minutti, Juan Vicente Alejandre Loya, Félix Emmanuel Pérez Talavera, Freddy José Ávila Cervera, Adriana Velazquez Burciaga, Oscar Morado Aramburo, Luis Alberto Piña Olguín, Adrian Soto-Rodríguez, Andrés Castañeda Prado, Patricio Santillán Doherty, Juan O Galindo Galindo, Luis Alberto Guízar García, Daniel Hernández Gordillo, Juan Gutiérrez Mejía

## Abstract

**- Importance:** Many COVID-19 prognostic factors for disease severity have been identified and many scores have already been proposed to predict death and other outcomes. However, hospitals in developing countries often cannot measure some of the variables that have been reported as useful.

**- Objective:** To assess the sensitivity, specificity, and predictive values of the novel LOW-HARM score (*L*ymphopenia, *O*xygen saturation, *W*hite blood cells, *H*ypertension, *A*ge, *R*enal injury, and *M*yocardial injury).

**- Design:** The score was designed using data from already published cohorts of patients diagnosed with COVID-19. Afterwards, it was calculated it in 438 consecutive hospital admissions at twelve different institutions in ten different cities in Mexico.

**- Setting:** Twelve hospitals in ten different cities in Mexico.

**- Participants:** Data from 438 patients was collected. Data from 400 patients (200 deaths and 200 survivors) was included in the analysis.

**- Exposure:** All patients had an infection with SARS-CoV-2 confirmed by PCR.

**- Main Outcome:** The sensitivity, specificity, and predictive values of different cut-offs of the LOW-HARM score to predict death.

**- Results:** Mean scores at admission and their distributions were significantly lower in patients who were discharged compared to those who died during their hospitalization 10 (SD: 17) vs 70 (SD: 28). The overall AUC of the model was 95%. A cut-off > 65 points had a specificity of 98% and a positive predictive value of 96%. More than a third of the cases (36%) in the sample had a LOW-HARM score > 65 points.

**- Conclusions and relevance:** The LOW-HARM score measured at admission is highly specific and useful for predicting mortality. It is easy to calculate and can be updated with individual clinical progression.

**KEY POINTS:** *Question:* Is it possible to predict mortality in patients diagnosed with COVID-19 using easy-to-access and easy-to-measure variables?

*Findings:* The LOW-HARM score (*L*ymphopenia, *O*xygen saturation, *W*hite blood cells, *H*ypertension, *A*ge, *R*enal injury, and *M*yocardial injury) is a one-hundred-point score that, when measured at admission, had an overall AUC of 95% for predicting mortality. A cut-off of ≥ 65 points had a specificity of 98% and a positive predictive value of 96%.

*Meaning:* The LOW-HARM score measured at admission is highly specific and useful for predicting mortality in patients diagnosed with COVID-19. In our sample, more than a third of patients met the proposed cut-off.

## INTRODUCTION

Recently, multiple prognostic factors for disease severity in patients diagnosed with COVID-19 have been identified^1-3^. In this regard, many prognostic scores have already been put forward to predict the risk of death and other outcomes (e.g. CALL score, ABC GOALS, Neutrophil-Lymphocyte index, etc.)^4–6^. However, hospitals in developing countries often cannot measure some of the variables included in these scores (D-dimer, ferritin, CT-scans, etc.).

Conversely, implementation of many of these scores is hampered by inclusion of subjective variables such as breathlessness^5^, data on pre-existing comorbidities^7^ (making it impossible to reassess prognosis according to the patients’ clinical evolution) or rely on cut-off values that are infrequently met by patients with COVID-19 in real-world settings.

Even further, developing countries have a lower number of critical-care beds^8^ and specialists per 100 000 people^9^. Thus, estimating mortality is essential for optimal resource allocation. It is not an understatement to say that, particularly in these resource-limited settings, improving these processes could save lives.

Prediction tools also have ethical applications and implications. Some triage systems repurpose scores to predict mortality in critical care patients, such as the SOFA (Sequential Organ Failure Assessment) score, as part of their decision framework^10,11^. However, there is compelling evidence highlighting the importance of generating and using disease-specific prediction tools or models in pandemic contexts^12^.

Furthermore, having context-specific predictive accuracy is essential for assisting the decision-making process in these extraordinary situations, for objectively tracking clinical status, and for providing realistic and accurate information to patients and their families about prognosis.

Mathematical models for estimating new cases of COVID-19 in the post-pandemic period agree there will be more than one “wave” of infections^13^, and serological surveys for estimating the dynamics of a population’s susceptibility, level of exposure and immunity to the virus support these predictions^14-16^. Therefore, an effective prognostic tool is still relevant even if most countries are already flattening their daily confirmed cases curve^17^.

The novel LOW-HARM score (*L*ymphopenia, *O*xygen saturation, *W*hite blood cells, *H*ypertension, *A*ge, *R*enal injury, and *M*yocardial injury) is a one-hundred-point score incorporating objective and easy-to-obtain data that are already measured as part of the clinical care of patients diagnosed with COVID-19. Moreover, its calculation is quick, reproducible, and easy to automate using free and intuitive, digital tools.

## METHODS

### Identification and selection of predictors

After thoroughly reviewing the available literature regarding prognostic factors for mortality in patients diagnosed with COVID-19, we chose epidemiological and clinical variables that allowed calculations of a likelihood-ratio (LR) and that were routinely available to most hospitals in developing countries^1-3^.

### Score design

Using the reported probability of death by age group ^18^ as the pre-test probability for death, the calculation for the LOW-HARM score is structured as follows:

1. Odds pre-test = Probability pre-test / (1-Probability pre-test).
2. Odds post-test = (Odds pre-test) x (LR SpO_2_) x (LR Diagnosis of Hypertension) x (LR elevation of cardiac enzymes) x (LR white blood cell count > 10 000 cells/mm^3^) x (LR total lymphocyte count < 0.8 cells/ mm^3^) x (LR serum creatinine >1.5 mg/dL).
3. Post-test probability = Post-test Odds / (1 + Post-test Odds).

Variables were dichotomised as present or absent. For oxygen saturation (SpO_2_), the cut-off was <88%; for cardiac enzymes, the cut-off was either a value >99^th^ percentile by each lab for high-sensitivity troponin, or > 185 U/L for creatine phosphokinase (CPK) or > 100 ng/mL for myoglobin.Pre-test odds for death for each age group were considered as follows: < 40 = 0.002, 40 - 49 = 0.004, 50 - 59 = 0.013, 60 - 69 = 0. 037, 70 −79 = 0.087, >80 = 0.174.

The calculated LRs were oxygen saturation < 88% = 6.85, previous diagnosis of hypertension = 2.06, elevated troponin. myoglobin or CPK = 6.00, leukocyte counts > 10 000 cells/μL= 4.23, lymphocyte counts < 800 cells/μL (<0.8 cells/mm3) = 2.89, serum creatinine > 1.5 mg/dL = 4.27.

### Sample size rationale

Considering that official estimations expect at least 10,000 critically-ill patients during this pandemic^19^, according to the formula for calculating samples from finite populations: n = N*X / (X + N - 1), where, X = Zα/22 -*p*(1 -p) / MOE, Zα/2 is 1.96, MOE is the margin of error, p is the sample proportion, and N is the population size, data from 385 patients are required to produce a statistically representative sample with an alpha of 0.05%.

### Analysis

Mean and dispersion values for the final scores and demographic variables were calculated. Sensitivity, Specificity, and predictive values for different score cut-off values were calculated. Areas under the Receiver Operating Characteristic curves (AUROC) were calculated for the whole model and for sensitivity and specificity using Simpson’s Rule. Two-tailed Student’s *t* tests for independent samples were used for comparing means, categorical variables were compared using a Xi^2^ test. A p-value of <0.05 for inferring statistical significance was used. All calculations were performed using Microsoft Excel® and STATA® v12 software.

This project and analysis strategy were pre-registered at the Open Science Framework (Code: 68er7) on April 21^st^, 2020. The original acronym for this score was changed to avoid confusion with the pre-existing CALL score. Data and a pre-print version of this document was made available at medRxiv.

### Data collection

Data was collected retrospectively (between April 30^th^ and May 20th) using an online formulary. Laboratory results and SpO_2_ values correspond to measurements at admission. Patients were grouped and analysed according to their clinical outcome (death vs survival).

### Data exclusion

We excluded from analysis all patients without a documented clinical outcome (e.g. transferred to another hospital for voluntary discharge), without complete clinical data or without a confirmatory PCR for the SARS-CoV-2 virus.

### Ethical approval

This study was assessed and approved by the Ethics Committee of the Instituto Nacional de Ciencias Médicas y Nutrición Salvador Zubirán on April 29^th^, 2020 (Reg. No. DMC-3369-20-20-1). Clinical data was anonymized, and its collection followed Good Clinical Practice Standards. This manuscript was written following EQUATOR reporting guidelines and the TRIPOD statement recommendations.

## RESULTS

### Demographic data

We obtained data from 438 patients treated at twelve hospitals in ten cities in Mexico: Instituto Nacional de Ciencias Médicas y Nutrición Salvador Zubirán, Instituto Nacional de Enfermedades Respiratorias, Centro Médico Nacional Siglo XXI, Centro Médico Nacional Occidente, Hospital Regional de Alta Especialidad de la Península de Yucatán, Hospital Regional de Alta Especialidad del Bajío, Hospital de la Beneficencia Española San Luis Potosí, Sistema de Salud Christus Muguerza (Puebla, Coahuila, Chihuahua, Nuevo León and Yucatán). A total of 38 patients were excluded according to the criteria previously mentioned. Table 1 summarises demographic characteristics of patients included in our analysis according to their clinical outcome.

**Table 1.** Demographic data according to clinical outcome.

| Variable | Survivors (n=200) | Deaths (n=200) | P value* |
| --- | --- | --- | --- |
| **Sex (%)** |  |  |  |
| Female | 67 (33.5) | 53 (26.5) | 0.127 |
| Male | 133 (66.5) | 147 (73.5) |  |
| **Age group, years (%)** |  |  |  |
| 20-29 | 18 (9) | 2 (1) | <0.0001 |
| 30-39 | 40 (20) | 14 (7) |  |
| 40-49 | 43 (21.5) | 28 (14) |  |
| 50-59 | 54 (27) | 57 (28.5) |  |
| 60-69 | 28 (14) | 54 (27) |  |
| 70-79 | 16 (8) | 33 (16.5) |  |
| >80 | 1 (0.5) | 12 (6) |  |
| **Weight, kg (SD)** | 80 (15.0) | 80.2 (17.2) | 0.800 |
| **Height, cm (SD)** | 168 (9) | 164.8 (9.2) | <0.0001 |
| **Body-mass index (SD)** | 28 (5) | 29.5 (5.8) | 0.013 |
| **Obesity (BMI ≥ 30 kg/m<sup>2</sup>) (%)** | 54 (27) | 78 (39) | 0.011 |
| **Required IMV (%)** | 22 (11) | 123 (61.5) | <0.0001 |
| **Length-of-stay, days (SD)** | 10 (7) | 8.1 (7.3) | 0.008 |
| **Diabetes Mellitus (%)** | 46 (23) | 78 (36) | 0.002 |
| **Pregnancy (%)** | 2 (1) | 0 (0) | 0.156 |
| **Smoking (%)** | 24 (12) | 25 (12.5) | 0.758 |
| **Immunocompromised (%)** | 9 (4.5) | 13 (6.5) | 0.380 |
| **COPD (%)** | 3 (1.5) | 35 (17.5) | <0.0001 |
| **CKD (%)** | 5 (2.5) | 8 (4) | 0.586 |
| **CAD (%)** | 3 (1.5) | 8 (4) | 0.126 |

\* Categorical variables were compared using a $X^2$ test, continuous variables were compared using an unpaired Student's *t* test. SD: standard deviation, IMV: invasive mechanical ventilation, COPD: chronic obstructive pulmonary disease, CKD: chronic kidney disease, CAD: coronary artery disease.

### Mortality score

The mean LOW-HARM scores were 10 (SD: 17) for those who survived their hospitalisation and 70 (SD: 28) for those who died (Mean difference: 61.1, 95% CI 56.7 - 65.6). **Figure 1** shows the score distribution for each outcome. **Table 2** summarizes the frequency of each score component in both groups.

**Figure 1.**
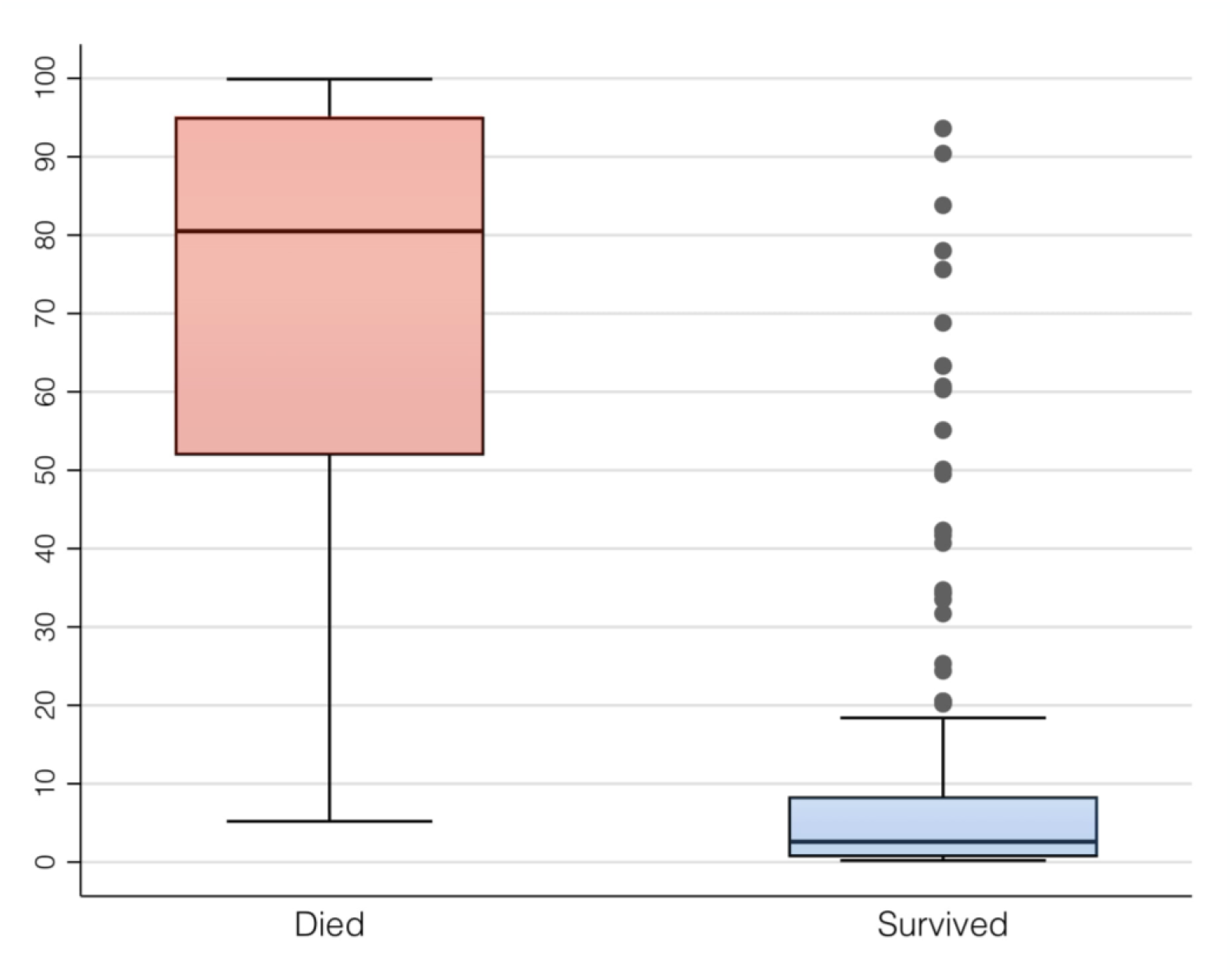
Distribution of scores in both groups.

**Table 2.** Frequency of each score component according to clinical outcome.

| **Variable** | **Survivors (n=200)** | **Deaths (n=200)** | **P value\*** |
| --- | --- | --- | --- |
| **Lymphocytes <800 cells/ μL** | 65 (32.5) | 146 (73) | <0.0001 |
| **SpO<sub>2</sub> <88% (%)** | 73 (36.5) | 191 (95.5) | <0.0001 |
| **White blood cells >10,000 cells/μL** | 20 (10) | 113 (56.5) | <0.0001 |
| **Hypertension (%)** | 40 (20) | 95 (47.5) | <0.0001 |
| **Serum creatinine >1.5 mg/dL (%)** | 4 (2) | 78 (36) | <0.0001 |
| **Cardiac Injury\* (%)** | 22 (11) | 118 (59) | <0.0001 |
| **CPK >185 U/L** | 16 (8) | 38 (19) |  |
| **hsTpi >99<sup>th</sup> P100** | 5 (2.5) | 78 (39) |  |
| **Myoglobin** | 1 (0.5) | 2 (1) |  |

*SpO<sub>2</sub>: peripheral capillary oxygen saturation*
*\* Categorical variables were compared using a $\chi^2$ test.*
*\* Cardiac injury was defined as elevation of high-sensitivity troponin I >99<sup>th</sup>*
*percentile, creatine phosphokinase serum levels >185 U/L or serum myoglobin levels >100 ng/mL.*

### Sensitivity, Specificity and Predictive Values

Sensitivity, Specificity, positive and negative predictive values, and AUROCs were calculated for cut-offs every five points. **Table 3** summarises these results.

**Table 3.** Sensitivity, Specificity, Positive and Negative Predictive Values, and AUROCs for different score cut-offs.

| **Cut-off** | **Se, %** | **Sp, %** | **PPV, %** | **NPV, %** | **AUROC (95%CI)** |
| --- | --- | --- | --- | --- | --- |
| **0** | 100 | 0 | 0 | 100 | - |
| **5** | 100 | 64 | 73.5 | 100 | 0.82 (0.79-0.85) |
| **10** | 97.5 | 78 | 82 | 97 | 0.88 (0.85-0.91) |
| **15** | 95 | 80.5 | 83 | 94 | 0.88 (0.85-0.91) |
| **20** | 92 | 84.5 | 85.5 | 91 | 0.88 (0.85-0.91) |
| **25** | 91 | 88.5 | 89 | 91 | 0.90 (0.87-0.93) |
| **30** | 86.5 | 89 | 89 | 87 | 0.88 (0.85-0.91) |
| **35** | 82.5 | 91.5 | 91 | 84 | 0.87 (0.84-0.90) |
| **40** | 82.5 | 91.5 | 91 | 84 | 0.87 (0.84-0.90) |
| **45** | 76.5 | 93.5 | 92 | 80 | 0.85 (0.82-0.88) |
| **50** | 76.5 | 94 | 93 | 80 | 0.85 (0.82-0.88) |
| **55** | 71 | 94.5 | 93 | 76.5 | 0.83 (0.79-0.86) |
| **60** | 70 | 95 | 93 | 76 | 0.83 (0.79-0.86) |
| **65** | 64.5 | 97 | 95.5 | 73 | 0.81 (0.77-0.84) |
| **70** | 56.5 | 97.5 | 95.5 | 69 | 0.77 (0.73-0.81) |
| **75** | 55.5 | 97.5 | 95.5 | 67 | 0.77 (0.73-0.80) |
| **80** | 50.5 | 98.5 | 97 | 66.5 | 0.75 (0.71-0.78) |
| **85** | 44.5 | 99 | 97.5 | 64 | 0.72 (0.68-0.75) |
| **90** | 34 | 99 | 97 | 60 | 0.66 (0.63-0.69) |
| **95** | 23.5 | 100 | 100 | 56.5 | 0.61 (0.59-0.65) |
| **100** | 0 | 100 | 100 | 0 | - |

Se: sensitivity, Sp: specificity, PPV: positive predictive value, NPV: negative predictive value, AUROC: area under the receiver operating characteristic curve
\*Positivity defined as having a score above the cut-off value and dying.

Sensitivity and specificity for the model was evaluated using a Sensitivity vs Specificity ROC curve. **Figure 2** plots these findings.

**Figure 2.**
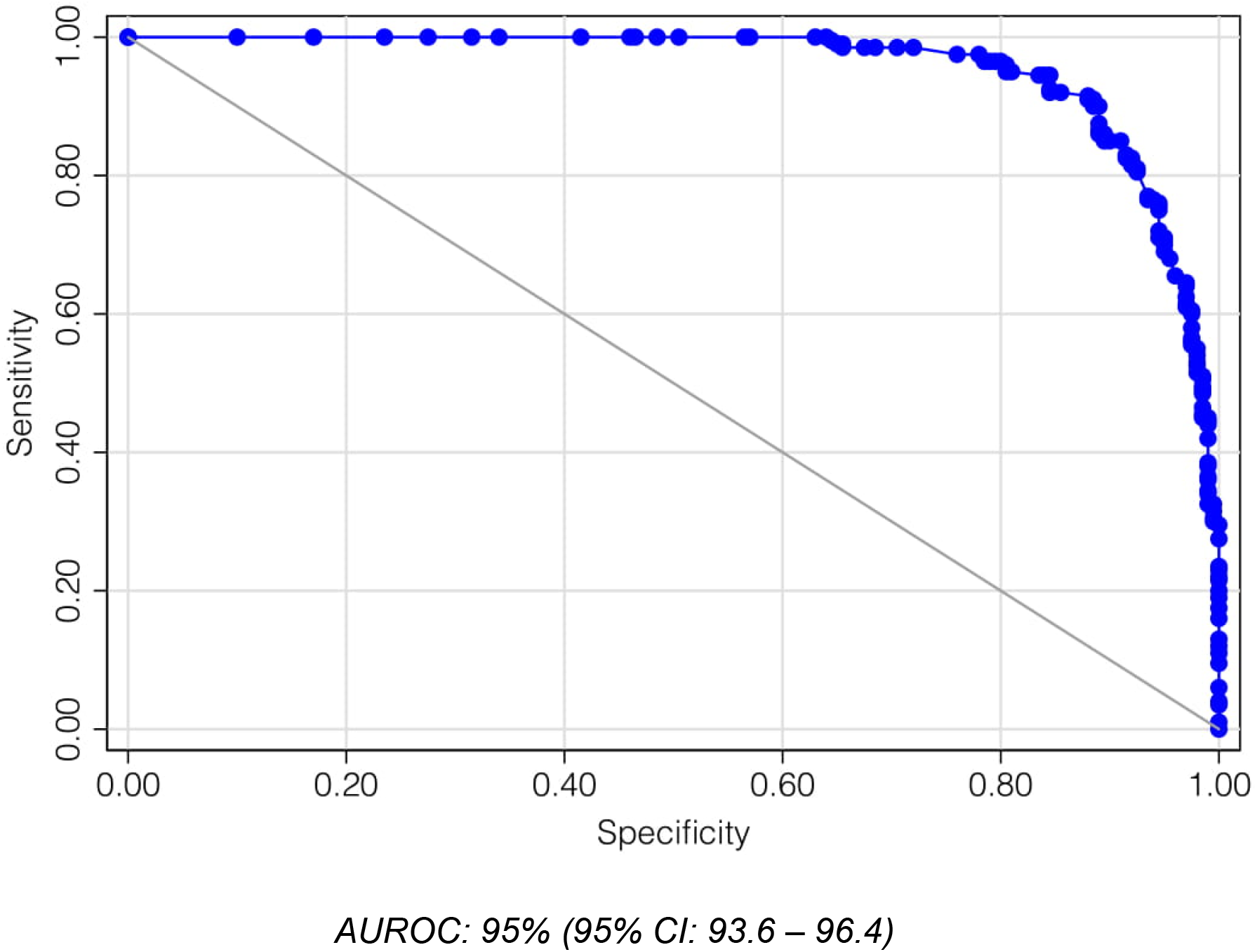
Sensitivity vs Specificity ROC curve and AUROC.

## DISCUSSION

Herein, we present the LOW-HARM score, a novel, easy-to-use and easy-to-measure tool to predict mortality in patients diagnosed with COVID-19. This score has been derived using prognostic factors observed specifically in cohorts of patients diagnosed with this disease^1-3^. Afterwards, we validated the LOW-HARM score using data from patients treated at twelve different institutions in ten different cities in a developing country.

Although no score or prediction tool should substitute clinical judgement, they can assist in many crucial aspects of clinical decision making. Accurately predicting which patients will not survive hospitalization can guide optimal resource allocation and support patients and clinicians in their decision-making process. To our knowledge, this is the first COVID-19-specific mortality prediction tool that is specifically designed to be used in low-income settings. However, it has other advantages that are relevant to more resourceful scenarios as well.

First, the LOW-HARM score allows dynamic reassessment and fine-tuning of its predictive capacities’ multiple times as the patient’s clinical condition evolves during hospitalization. Even in a resourceful environment, many of the variables used in other scores are not dynamic and/or are not measured frequently (comorbidities, CT-scans, IL-6, among others). Contrastingly, cardiac enzymes are repeatedly measured as an independent prognostic tool and complete blood counts and serum creatinine levels are measured almost daily. This allows to make comparisons and to update predictions as clinical evolution progresses.

Additionally, simply adding or subtracting comorbidities does not consider complicated aspects, such as how controlled were those diseases in the first place^7^. It would not be accurate to assume a 50-year-old patient living with diabetes without proper follow-up or treatment has the same prognosis as one who has successfully controlled their disease; similar principles would apply to other comorbidities such as cancer or immunosuppression, who have been shown to negatively impact prognosis in patients diagnosed with COVID-19.

There are potentially more useful or accurate time-points for measuring the LOW-HARM score (i.e. right before initiating mechanical ventilation or 24 hrs after initiating treatment). It may also be possible that the rate of change in the score, showing either improvement or worsening, has more prognostic value than its assessment at admission. Additionally, the fact that in the twenty-two patients who had positive cardiac injury biomarkers and survived, elevated CPK was three times more frequent than elevated troponin, suggests that prediction power could improve if cardiac enzymes are assigned values in the calculation. However, these comparisons are outside the scope of this study and will be pursued in following studies.

Finally, scores usually have practical and logistical limitations that preclude their applicability and adoption in real-world settings. As a part of this study, we have created a free digital tool where the calculation of the LOW-HARM score can be automatized, allowing quick, frequent (even daily), and reproducible predictions as the patient’s status evolves^20^.

### Choosing a cut-off

A numerical score is useful for making comparisons or tracking clinical evolution on their own, however, having a cut-off value could be useful for decision making.

The largest AUROC was observed using a cut-off score of 25 (0.90, 95% CI 0.87-0.93). However, when predicting mortality, and particularly when resources might be allocated based on this prediction, it is preferable to avoid false positive errors (predicting a patient will die when they will survive). Therefore, we propose 65 points as a more clinically useful cut-off, since it has a more than five times lower rate of false positive results if compared against the 25-point cut-off (2% vs 11 %).

It should be emphasized that, regardless of its diagnostic accuracy, proposing a score cut-off is as useful as the proportion of times this cut-off is met. In this case, 137/400 (36%) patients had a score above 65 which means it is possible to predict mortality with a specificity of 98% and a positive predictive value of 96% in more than a third of the patients at the time of admission.

It is also worth mentioning that, compared with other countries, Mexico has a high proportion of young people living with hypertension, diabetes, and obesity^21-23^. Therefore, results may vary in countries with a different demographic composition and burden of chronic disease.

Although data was consecutively obtained, our study has the inherent limitations of all retrospective studies. Prospective validation and research for comparing measurements at different time-points is warranted.

## CONCLUSION

The LOW-HARM score measured at the time of admission has a high accuracy in predicting mortality in patients diagnosed with COVID-19 requiring hospitalization. This score provides a disease- and context-specific tool that uses widely available and easily obtainable variables that promote generalized applicability in resource-limited settings. It can be calculated multiple times during patient follow-up using an intuitive and accessible online platform which allows individualized patient care.

## Data Availability

Anonymized data will be public.

https://docs.google.com/spreadsheets/d/1DH4nmfWi-T55Es-Y0CZYzXAwyibcVe_LQ7GyNwK9YWM/edit?usp=sharing

## ACKNOWLEDGEMENTS

Ricardo Sanginés, Dr Berenice Martínez Lara at Centro Médico Nacional de Occidente. All authors thank their respective institutions for their support.

## AUTHOR CONTRIBUTION

Adrian Soto-Mota designed the prediction score. Juan Gutiérrez Mejía, Braulio Alejandro Marfil-Garza and Adrian Soto-Rodríguez collaborated in designing and executing data analysis. Erick Martínez Rodríguez, José Omar Barreto Rodríguez, Alicia Estela López Romo, Paolo Alberti Minutti, Juan Vicente Alejandre Loya, Félix Emmanuel Pérez Talavera, Freddy José Ávila Cervera, Adriana Velazquez Burciaga, Oscar Morado Aramburo, Luis Alberto Piña Olguín, Andrés Castañeda Prado obtained clinical and demographic data. Patricio Santillán Doherty, Juan O Galindo, Luis Alberto Guízar García, Daniel Hernández Gordillo provided mentorship. All authors revised this manuscript and data analysis.

## FUNDING

This project received no external funding.

## CONFLICT OF INTERESTS

None of the authors declares financial interests or personal relationships that could have influenced the work reported in this study. ASM and BAMG are currently funded by the Instituto Nacional de Ciencias Médicas y Nutrición Salvador Zubirán.

## Notes

### Competing Interest Statement

The authors have declared no competing interest.

### Clinical Protocols

https://osf.io/qzunb

### Author Declarations

This study was assessed and approved by the Ethics Committee of the Instituto Nacional de Ciencias Medicas y Nutricion Salvador Zubiran on April 29th, 2020 (Reg. No. DMC-3369-20-20-1).

## REFERENCES

1. Ruan Q, Yang K, Wang W, Jiang L, Song J. Clinical predictors of mortality due to COVID-19 based on an analysis of data of 150 patients from Wuhan, China. Intensive Care Med. 2020. doi:10.1007/s00134-020-05991-x

2. Zhou F, Yu T, Du R, et al. Clinical course and risk factors for mortality of adult inpatients with COVID-19 in Wuhan, China: a retrospective cohort study. Lancet. 2020;395(10229):1054–1062. doi:10.1016/S0140-6736(20)30566-3

3. Xie J, Hungerford D, Chen H, et al. Development and external validation of a prognostic multivariable model on admission for hospitalized patients with COVID-19. *medRxiv*. 2020. doi:10.1101/2020.03.28.20045997

4. Ji D, Zhang D, Xu J, et al. Prediction for Progression Risk in Patients with COVID-19 Pneumonia: the CALL Score. *medRxiv*. 2020. doi:10.1093/cid/ciaa414/5818317

5. Mejía-Vilet JM, Córdova-Sánchez BM, Fernández-Camargo DA, et al. Derivation of a score to predict admission to intensive care unit in patients with covid-19: the abc-goals score. *medRxiv*. 2020. doi:10.1101/2020.05.12.20099416

6. Liu J, Liu Y, Xiang P, et al. Neutrophil-to-Lymphocyte Ratio Predicts Severe Illness Patients with 2019 Novel Coronavirus in the Early Stage. medR. 2020. doi:10.1101/2020.02.10.20021584

7. Bello-Chavolla OY, Bahena-Lopez JP, Antonio-Villa NE, et al. Predicting mortality attributable to SARS-CoV-2: A mechanistic score relating obesity and diabetes to COVID-19 outcomes in Mexico. *medRxiv*. May 2020:2020.04.20.20072223. doi:10.1101/2020.04.20.20072223

8. Acute care hospital beds per 100 000 - European Health Information Gateway. https://gateway.euro.who.int/en/indicators/hfa_478-5060-acute-care-hospital-beds-per-100-000/. Accessed May 25, 2020.

9. Heinze-Martin G, Olmedo-Canchola VH, Bazán-Miranda G, Bernard-Fuentes NA, Guízar-Sánchez DP. Los médicos especialistas en México. Gac Mex. 2018;154(3). doi:10.24875/GMM.18003770

10. White DB, Lo B. A Framework for Rationing Ventilators and Critical Care Beds During the COVID-19 Pandemic. JAMA. March 2020. doi:10.1001/jama.2020.5046

11. White DB, Katz MH, Luce JM, Lo B. Who should receive life support during a public health emergency? Using ethical principles to improve allocation decisions. Ann Intern Med. 2009;150(2):132–138. doi:10.7326/0003-4819-150-2-200901200-00011

12. Khan Z, Hulme J, Sherwood N. An assessment of the validity of SOFA score based triage in H1N1 critically ill patients during an influenza pandemic. Anaesthesia. 2009;64(12):1283–1288. doi:10.1111/j.1365-2044.2009.06135.x

13. Kissler SM, Tedijanto C, Goldstein E, Grad YH, Lipsitch M. Projecting the transmission dynamics of SARS-CoV-2 through the postpandemic period. Science (80-). 2020;368(6493):eabb5793. doi:10.1126/science.abb5793

14. Doi A, Iwata K, Kuroda H, et al. Title: Seroprevalence of novel coronavirus disease (COVID-19) in Kobe, Japan. *medRxiv*. 2020. doi:10.1101/2020.04.26.20079822

15. Fontanet A, Tondeur L, Madec Y, et al. Cluster of COVID-19 in northern France: A retrospective closed cohort study. *medRxiv*. April 2020:2020.04.18.20071134. doi:10.1101/2020.04.18.20071134

16. Bendavid E, Mulaney B, Sood N, et al. COVID-19 Antibody Seroprevalence in Santa Clara County, California. *medRxiv*. April 2020:2020.04.14.20062463. doi:10.1101/2020.04.14.20062463

17. Coronavirus (COVID-19) Cases - Statistics and Research - Our World in Data. https://ourworldindata.org/covid-cases. Accessed May 25, 2020.

18. Team TNCPERE, Team TNCPERE. The Epidemiological Characteristics of an Outbreak of 2019 Novel Coronavirus Diseases (COVID-19) — China, 2020. China CDC Weekly, 2020, Vol 2, Issue 8, Pages 113-122. 2020;2(8):113–122. doi:10.46234/CCDCW2020.032

19. Versión estenográfica | Conferencia de prensa. Informe diario sobre coronavirus COVID-19 en México | Presidencia de la República | Gobierno | gob.mx. https://www.gob.mx/presidencia/articulos/version-estenografica-conferencia-de-prensa-informe-diario-sobre-coronavirus-covid-19-en-mexico-238145. Accessed May 24, 2020.

20. LowHarmScore COVID-19. https://lowharmcalc.com/. Accessed May 25, 2020.

21. Campos-Nonato I, Hernández-Barrera L, Flores-Coria A, Gómez-Álvarez E, Barquera S. Prevalencia, diagnóstico y control de hipertensión arterial en adultos mexicanos en condición de vulnerabilidad. Resultados de la Ensanut 100k. Salud Publica Mex. 2019;61(6, nov-dic):888. doi:10.21149/10574

22. Shamah-Levy T, Campos-Nonato I, Cuevas-Nasu L, et al. Sobrepeso y obesidad en población mexicana en condición de vulnerabilidad. Resultados de la Ensanut 100k. Salud Publica Mex. 2019;61(6, nov-dic):852. doi:10.21149/10585

23. Rojas-Martínez R, Basto-Abreu A, Aguilar-Salinas CA, Zárate-Rojas E, Villalpando S, Barrientos-Gutiérrez T. Prevalencia de diabetes por diagnóstico médico previo en México. Salud Publica Mex. 2018;60(3, may-jun):224. doi:10.21149/8566

